# Cortico-Pallidal Beta Dynamics Underlie Impaired Turning in Parkinson’s Disease

**DOI:** 10.1101/2025.11.25.25340849

**Authors:** Poojan D. Shukla, Jessica E. Bath, Kenneth H. Louie, Hamid Fekri Azgomi, Eleni Patelaki, Jacob Marks, Jannine Balakid, Doris D. Wang

## Abstract

Turning while walking is one of the most common yet complex human movements, and it is frequently impaired in Parkinson’s disease (PD), leading to falls and loss of independence. The neural mechanisms underlying this difficulty remain unclear. Using chronically implanted devices that simultaneously record and stimulate the brain, we measured activity in the motor cortex and basal ganglia of individuals with PD during natural walking and turning. Successful turns were marked by reduced beta-band activity and flexible communication between cortical and pallidal regions, whereas impaired turns showed excessive beta synchrony that rigidly constrained movement. Medication and deep brain stimulation improved turning through distinct circuit mechanisms, dopamine suppressing abnormal pallidal beta activity and its communication with the cortex, and stimulation restoring cortical flexibility. These findings reveal how dynamic cortical–basal ganglia interactions enable complex movement and establish circuit targets for adaptive brain stimulation to reduce falls in PD.

## Introduction

Axial motor deficits, including impairments in gait, balance, and turning, are among the most disabling symptoms of Parkinson’s disease (PD).^1^ Because axial motor behaviors are central to daily function, difficulty performing them disproportionately reduces quality of life. Turning is a key axial behavior and a fundamental component of human gait. On average, individuals perform over 700 turns per day, and 35-45% of daily steps involve turning during routine activities.^2,3^ In PD, however, turning is frequently impaired as greater than 50% of patients with PD report difficulty turning.^4,5^ Compared to healthy individuals, patients with PD exhibit slower turning speeds, increased step counts, reduced turning angles and arcs, and greater variability across these metrics.^3,5–10^ Moreover, turns in PD are often less stable when matched for speed with healthy subjects and are typically described as “en-bloc”, a pattern marked by impaired axial rotation that prevents segmental rotation of the head, shoulders, and pelvis in a smooth fashion.^9,11^ Freezing of gait, characterized by the sudden and brief inability to step, is also more prevalent during turns.^8,10,12^ Collectively, these difficulties substantially increase fall risk and contribute to the overall burden of axial disability.^6,8,9,11–14^

Pharmacological and surgical treatments for PD, including levodopa and deep brain stimulation (DBS), have demonstrated variable and often unpredictable effects on axial symptoms such as turning. While dopamine replacement therapy can improve gait and turning metrics in some patients, such as speed and stride length, it may paradoxically exacerbate postural instability and impair balance in some individuals.^1,15–18^ Similarly, axial behaviors such as balance and posture may improve, worsen, or remain unchanged following DBS.^1,14,19–21^ These inconsistent therapeutic responses are further complicated by the absence of standardized tools to evaluate turning quality. For instance, the postural instability and gait difficulty (PIGD) score of the MDS-UPDRS III does not specially assess turning, and other clinical assessments of balance such as the Mini-BEST test, offer only coarse ratings of turn performance.^1,22^ As a result, existing clinical rating scales may lack the resolution necessary to capture the full spectrum of turning strategies or to detect meaningful changes across individuals. ^6,23^

Despite the ubiquity and clinical relevance of turning during gait, the neurophysiological mechanisms underlying this behavior remains poorly understood. Turning requires the seamless integration of sensory inputs, motor planning, and dynamic postural adjustments, relying on coordinated activity across cortical and subcortical structures, including the basal ganglia, cerebellum, and sensorimotor cortices.^24–27^ Impaired turning in PD may result from inadequate cortical compensation for basal ganglia dysfunction, but direct neurophysiological evidence is limited.

To address these gaps, we analyzed multisite intracranial recordings from individuals with PD using an investigational bidirectional neural interface during an overground walking task that included straight-ahead walking and 180° turns (Figure 1). We first characterized how turning-related neural signatures differ from those observed during straight walking in both the globus pallidus internus (GPi) and cortex. We then introduced and validated a novel metric of turning quality, the Turning Performance Index (TPI), to assess the heterogeneous effects of levodopa and DBS on turning behavior. Finally, we examined the neurophysiological correlates of improved versus unimproved turning across treatment conditions and trained machine learning models to classify turning periods from neural recordings and predict turn quality. These findings advance our understanding of turning impairment in PD and point to modifiable neurophysiologic mechanisms that may underlie treatment responses. This work provides a foundation for developing targeted neuromodulation strategies to improve turning and other axial motor symptoms in PD.

**Figure 1.**
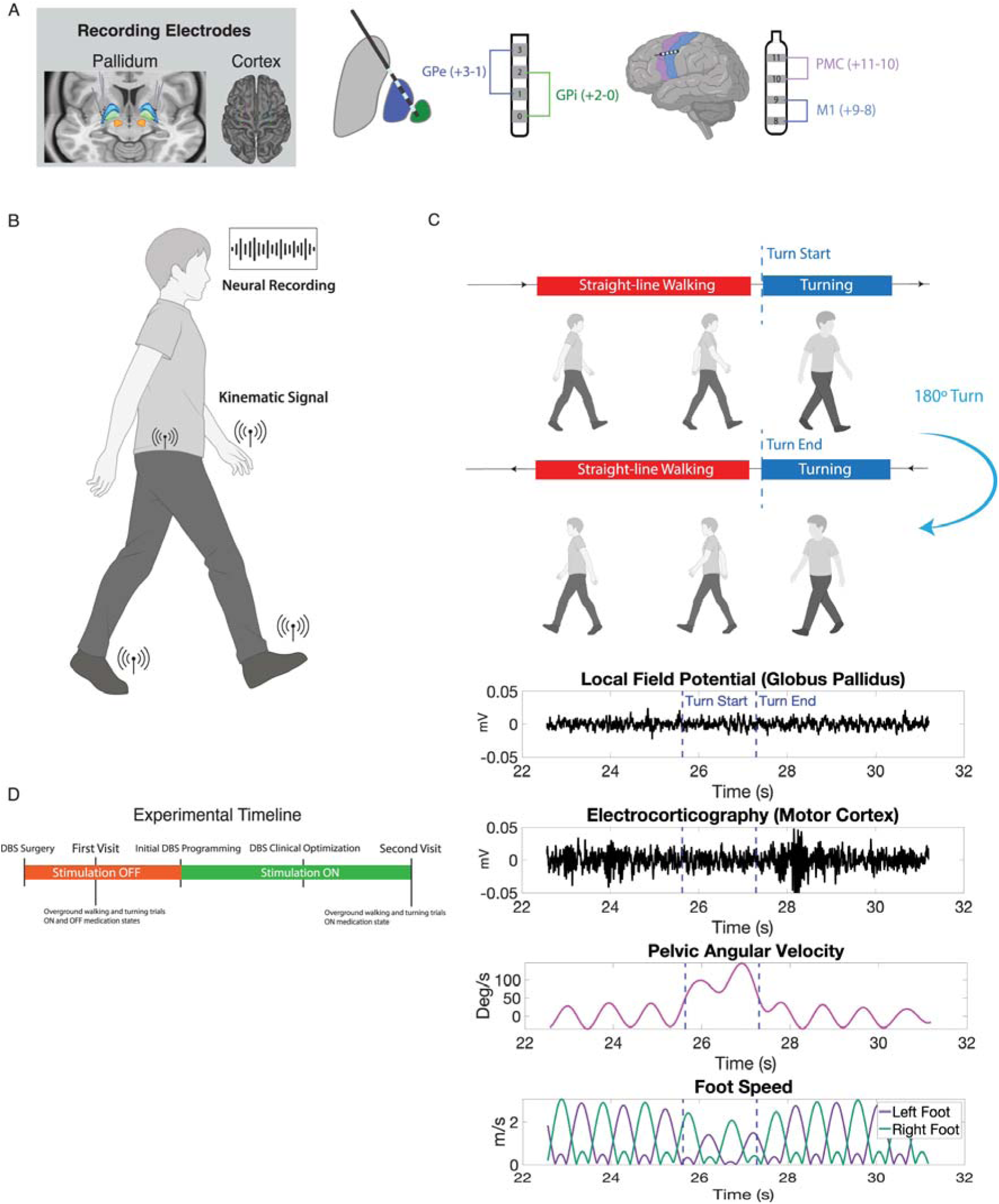
Experimental overview and workflow. (**A**) Implanted bidirectional neural interface facilitates neural recording from the pallidum and cortex. **(B)** Kinematic signals are recorded from body worn IMU sensors during overground walking task. (**C**) Overview of walking task. Subjects completed trials of walking in a loop featuring 180 degree turns. Direction of turning was prespecified. Also shown are traces of pelvic angular velocity and foot speed which are recorded from body worn IMUs as well as time series of local field potential and electrocorticography recordings. (**D**) Sample timeline of stimulation ON and OFF testing along with medication states followed by each patient.

## Materials and methods

### Patient selection, surgical procedure, and electrode localization

Five patients with idiopathic PD (three male, two females age range 57-71, Supplementary Table 1) underwent implantation of a bidirectional neural interface (Summit RC+S, Medtronic Inc.) as part of an ongoing clinical trial (ClinicalTrials.gov ID: NCT-0358289) with IRB approval (UCSF IRB# 20-32847). Inclusion criteria were surgical candidacy for DBS, baseline MoCA (Montreal Cognitive Assessment) score of at least 21, and disease duration of at least four years.

The device streamed local field potentials (LFPs) from the globus pallidus internus (GPi) and externus (GPe) and recorded electrocorticography (ECoG) potentials from primary motor (M1) and premotor cortices (PMC) (Figure 1 A-C). Electrode localization followed established pipelines.^28–36^ Four patients received bilateral implants; subject 5 underwent left-sided unilateral implantation. For subject 2, only left-sided data were analyzed due to a staged right implantation 10 months later.

### Experimental setup, data collection, and data alignment

#### Experimental setup

Subjects performed multiple trials of an overground walking task consisting of approximately 200 non-turning steps in a 6-meter loop. At each end, patients executed 180° turns to the left (counterclockwise) or right (clockwise) as instructed before the trial start. Walking tasks were completed in the ON and OFF medication states prior to DBS activation. The ON and OFF medication (ON/OFF med) states were defined as previously described. ^29^ Briefly, ON med was defined as 30 minutes following the last dose, and OFF med as withholding one or more doses, as tolerated. Patients then performed the walking tasks after clinical DBS optimization in both medication states (if tolerated, Figure 1D).

#### Data collection and alignment

During all trials, surface EMG (Tirgno, Delsys Inc., Natik, MA) and inertial measurement units (IMU, Xsens, Movella, Netherlands) collected biomechanical measurements. Force-sensitive resistors (FSRs) were placed under the calcaneus, hallux, and first and fifth metatarsal heads to capture gait events. Neural data were continuously streamed at 500 Hz from GPi and GPe contact pairs (0–2, 1–3) and from adjacent ECoG electrodes over M1 and PMC. Data were processed using open-source pipelines (https://github.com/openmind-consortium/Analysis-rcs-data) as described previously.^28–30,36,37^

Neural, EMG, and IMU signals were aligned in MATLAB (Mathworks, Inc., Natick, MA, version 2024b) by synchronizing peaks from the on-board accelerometer, IMU, and EMG sensors, following previously described methods.^28–30,36,38^

#### Kinematic Signal Processing and Turn Identification

All kinematic signals were sampled at 60 Hz and were low-pass filtered in MATLAB using a 1.5 Hz 4^th^ order Butterworth filter. Turns were identified using patient-specific pelvic angular velocity thresholds of 30-70 degrees/second, as validated previously.^7^ Straight-line walking epochs were defined as all steps outside turning epochs, excluding the stride immediately before and after each turn.

### Definition and validation of the TPI

#### Turn kinematic parameter extraction

Kinematic parameters associated with turning performance were identified from literature search.^5–11,13,39–41^ A correlation matrix of all variables is shown in Figure 3A. Stability was defined following published descriptions.^13,42–44^ Briefly, the base of support (BOS) was bounded laterally by line segments through the positions of the foot and toe as reported by Xsens and anteriorly by lines perpendicular to these segments. The lateral and anterior-posterior (AP) margins of stability (MOS) were computed as the perpendicular distances from the center of mass (COM) to the corresponding BOS borders. The inverted pendulum model was used to derive extrapolated COM (ECOM) positions:

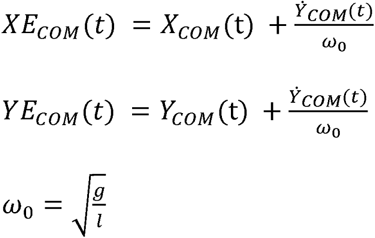

From these, we calculated COM and ECOM percent stable and minimum MOS in both lateral and AP directions. Percent stable reflected the proportion of the turn during which the COM or ECOM remained within the BOS (Fig. 3D). Minimum MOS represented the smaller of left/right MOS values. For AP MOS, only the leading foot was considered.

#### TPI construction

Principal component analysis (PCA) identified kinematic variables most correlated with dominant axes of variation, guiding feature selection for the TPI. A composite stability metric was defined as the mean of the four lateral stability measures (COM and ECOM minimum MOS, COM and ECOM percent stable). The final TPI was defined as

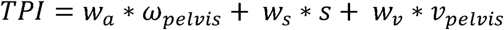

where *ω_pelvis_* represents pelvic angular velocity, *s* is composite stability, and ν*_pelvis_* is linear velocity measured at the pelvis.

#### TPI validation

TPI validity was assessed by correlating TPI values with blinded physical therapist ratings of 40 randomly selected turns across five subjects (ordinal scale 1–8). In addition, two reviewers evaluated turning performance overall across conditions within each subject and assigned ordinal ranks. A third reviewer broke any ties. Model coefficients were selected by optimizing Spearman correlation such that the absolute value of coefficients sum to one. To prevent overfitting, an additional optimization constraint was the overall TPI ratings of conditions agreed with the clinical assessment. The optimal weights found were:

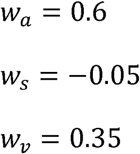

### Neural Signal Processing

#### Filtering and artifact rejection

LFPs were high-pass filtered at 2 Hz using a Butterworth filter. Stimulation artifacts were removed by notch filtering, and EKG contamination was eliminated via template subtraction when present.^45^ Channels referenced to the stimulating electrode were excluded from ON-stimulation analyses due to residual artifacts. High-frequency artifacts were detected using a custom algorithm.^38^ Signals were bandpass filtered from 75-150 Hz, transformed using multitaper spectral estimates, Z-normalized, and trials exceeding patient-specific z-score thresholds (30–80) were excluded.

#### Time-frequency analysis

PSD (power spectral density) was calculated using Welch’s method implemented through ‘pwelch’ in MATLAB (window size 250 samples, 90% overlap, 256 NFFT). For epochs shorter than 500ms, the largest window size smaller than the epoch length was used. Absolute power was computed using the STFT (short-time Fourier Transform) through the ‘spectrogram’ function with identical parameters and was also used to generate time-frequency spectrograms. Coherence was analyzed through wavelet coherence using ‘wcoherence’ (2-150 Hz, 32 voices per octave). Analyses focused on canonical frequency bands: theta (4-8 Hz), alpha (8.5-12 Hz), low beta (13-20 Hz), high beta (20.5-30 Hz), and low gamma (30.5-50 Hz).

#### Normalization

When comparing PSDs across multiple conditions and aggregating the results of all subjects, percent normalized power was defined as the power in each frequency bin divided by the total power from 2-50 Hz. Trials were normalized and pooled across hemispheres for bilaterally implanted patients.

#### Correlation analyses

Spearman’s correlation was conducted between kinematic variables and absolute power, normalized power, and beta burst features. For kinematic variables such as angular acceleration that were sampled throughout a turn, point-by-point correlation was performed with average power at that given time point. Otherwise, trial level turn metrics were correlated with power averaged across each trial.

#### Beta-bursting analysis

Filtered neural signals were first Z-normalized and bandpass filtered for the low beta, high beta, or entire beta band. Beta-bursts longer than 100ms in duration and exceeding the 75^th^ percentile of the signal’s amplitude envelope were considered for downstream analysis.^46–49^ Burst duration and amplitude were correlated with TPI while burst count was dichotomized into “low” and “high” TPI groups using the median split.

#### Machine learning analysis

Model training and assessment was performed using Python (version 3.9.21) and the scikit-learn library (version 1.1.3). LDA (linear discriminant analysis) classifiers were trained individually on subjects. First, all straight-line walking and turning epochs were divided into non-overlapping two second windows, with smaller windows permitted for shorter epochs. For each brain region, average power in the canonical frequency bands and in each possible frequency band from 2-50Hz was extracted in addition to average coherence across regions for these bands, yielding 4401 features per subject.

A random forest classifier (n=1000 trees, all variables sampled at each split) was trained using 5-fold CV (cross validation) to rank feature importance. The top 20 features were selected to train an LDA classifier using 5-fold CV on the entire training dataset. The cross-validation AUC was reported as the final AUC. To predict TPI, a random forest regressor trained using 5-fold CV or order features by mean importance. Nested CV was performed with five different model architectures: random forest regression, gradient boosted regression, LASSO, Elastic Net, and ridge regression. The outer CV loop was used to optimize model hyperparameters and the inner CV loop was used to select the model that achieved the highest mean R^2^ after 5-fold CV, which was reported as the final model R^2^.

### Statistical analysis

Continuous variables were compared using two-sided Wilcoxon rank-sum tests with Benjamini-Hochberg correction, implemented using the ‘fdr_bh’ function in MATLAB. Adjustment for multiple comparisons was applied to all comparisons made per channel, and all p values reported have been adjusted.

Classifier AUC significance was assessed by 1000-iteration permutation testing, comparing model AUC to the null distribution of permuted labels.

## Results

### Cortical beta desynchronization distinguishes turning from straight-line walking

We first explored differences in neural biomarkers between turning and straight-line overground walking epochs in both ON and OFF med states, prior to DBS activation. Across five subjects and eight implanted hemispheres, we analyzed 394 turning trials with 399 straight-line walking epochs. Compared to straight-line walking (Str), both M1 and PMC exhibited decreased normalized low beta (turning vs Str: 3.37% vs 3.77%, *p=*0.0028) and high beta power (turning vs Str: 3.64% vs 3.93%, *p*=0.0087) during turning (Figure 2 A, B). GPe also exhibited diminished high beta power during turning compared to straight-line walking (turning vs Str: 1.77% vs 1.97%, *p=*0.0061). No significant spectral differences were seen in the GPi. Subject specific changes in power and coherence are shown in Supplementary Figure 1.

**Figure 2.**
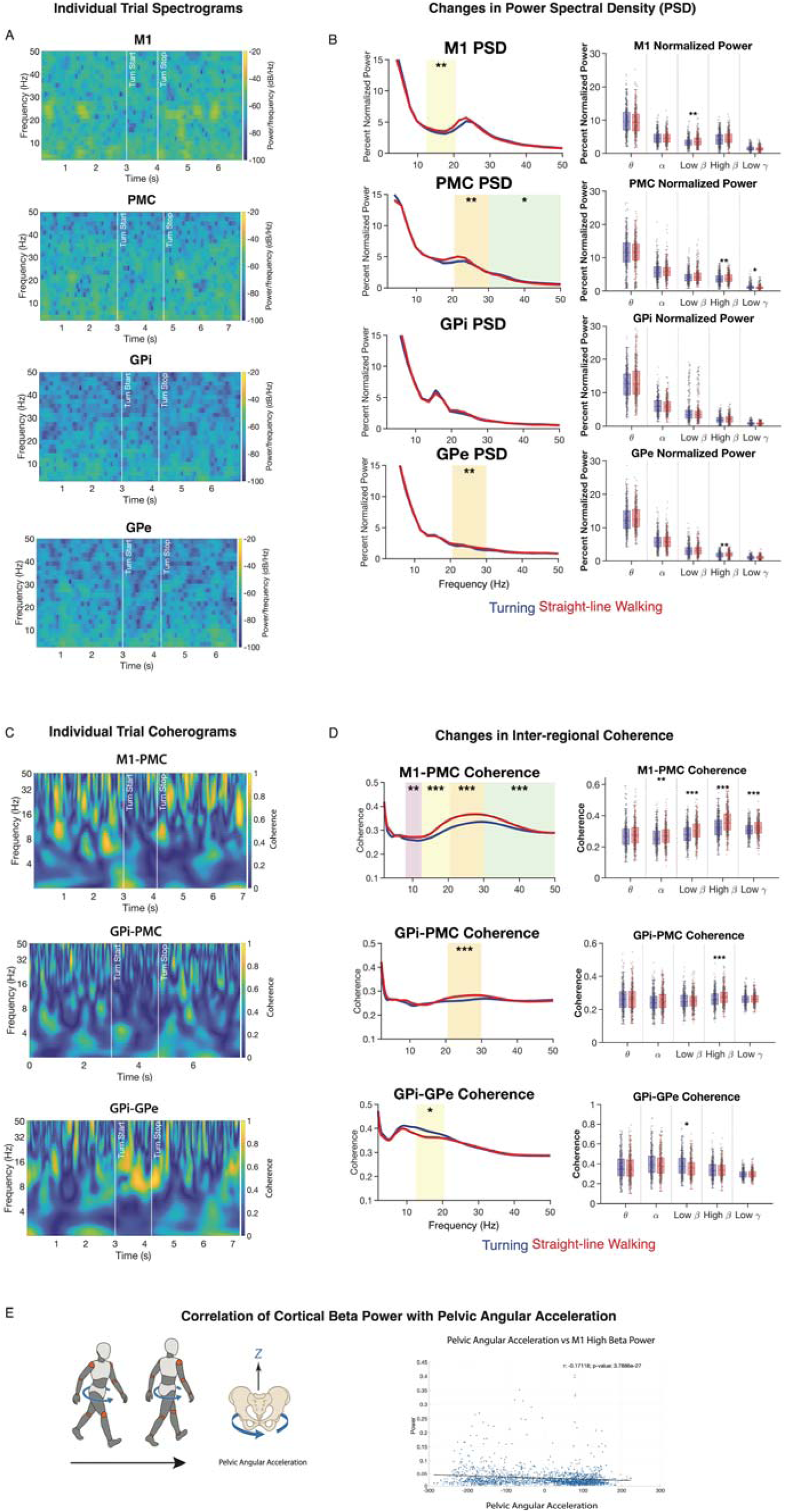
Neurophysiology of straight-line walking versus turning. **(A**) Spectrograms of individual turning trials and surrounding straight-line walking periods for globus pallidus internus (GPi), globus pallidus externus (GPe), primary motor (M1), and premotor (PMC) cortices. No clear differences across epochs are easily seen in the GPi while the GPe, M1, and PMC spectrograms better illustrate a decrease in beta power from “Turn Start” to “Turn Stop.” (**B**) Plot of normalized power spectral density averaged across all trials for straight-line walking and turning, along with box and whisker plot showing distribution of power in canonical frequency bands. All data comes from DBS OFF trials, with channels for the GPi, GPe, M1, and PMC shown. **(C**) Coherograms of individual turning trials and surrounding straight-line walking periods for GPi-GPe, GPi-PMC, and M1-PMC coherence. (**D**) Inter-regional coherence during straight-line walking and turning, along with corresponding box and whisker plots are shown for the GPi-GPe, GPi-PMC, and M1-PMC. (**E**) Schematic illustration of pelvic angular acceleration during turning and correlation between M1 high beta power and pelvic angular acceleration for one subject. For all panels, * indicates *p* <0.05, ** indicates *p*<0.01, and *** indicates *p*<0.001. Shaded colored regions in spectral power and coherence plots indicate a significant difference in the specified frequency band (theta 4-8 Hz, alpha 8.5-12 Hz, low beta 13-20 Hz, high beta 20.5-30 Hz, and low gamma 30.5-50 Hz).

**Figure 3.**
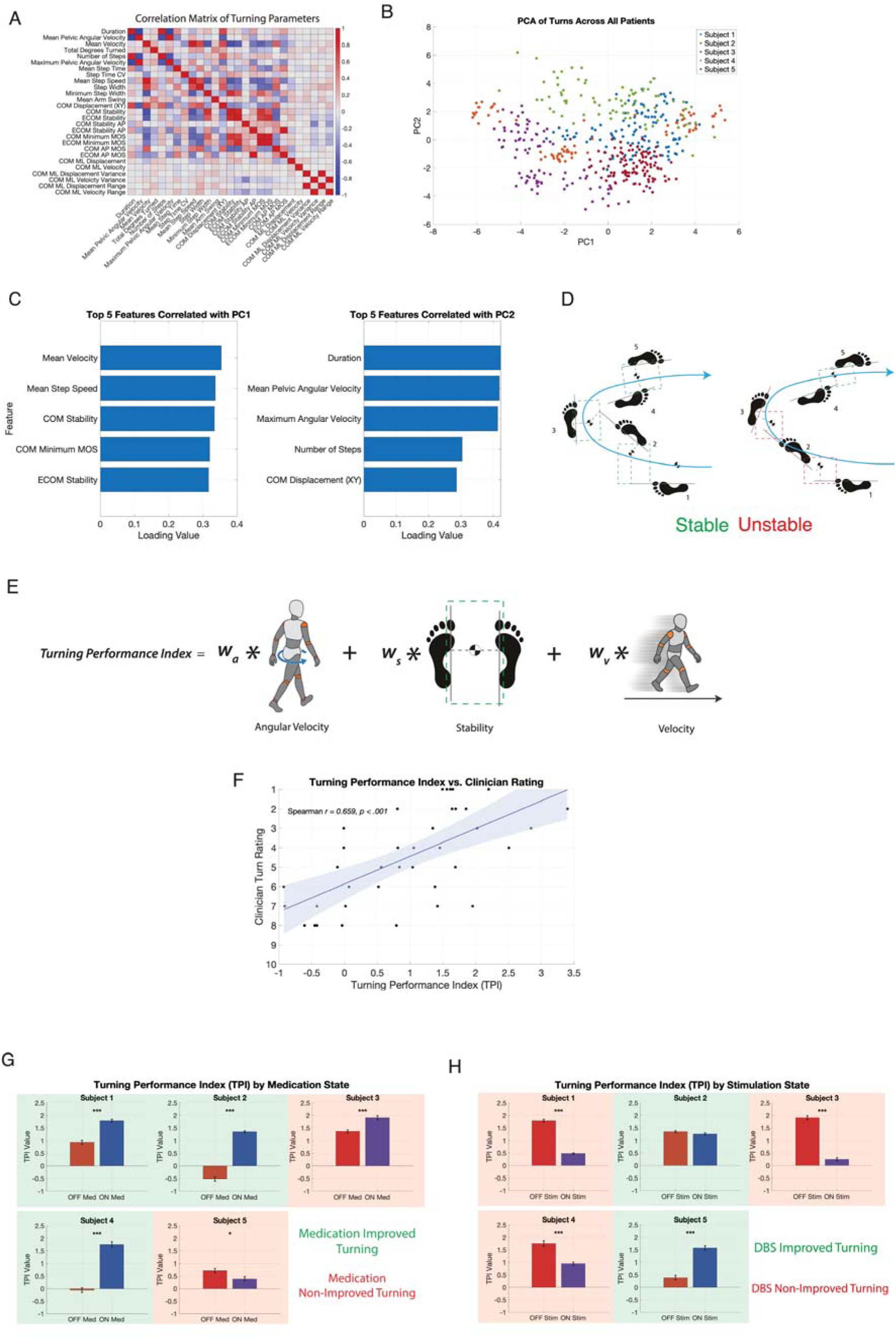
Definition and validation of a turning performance index. (**A**) Correlation matrix showing correlation between all turning parameters analyzed. (**B**) Principal component analysis of all turns analyzed based on their turning parameters. (**C**) Top five turning parameters most correlated with the first and second principal component by absolute value of loading coefficient. (**D**) Illustration of the stability measurement. Checkered circle indicates center of mass, dashed grey lines indicate distance from center of mass to lateral edges of base of support. Green boxes indicate instances of the center of mass staying within the base of support while red boxes denote periods when the center of mass lies outside, resulting in instability. (**E**) Schematic illustrating the final turning performance index formulation. (**F**) Validation of our turning performance index with clinician rating. (**G**) Turning performance index applied to all subjects in stimulation off trials. Green boxes highlight subjects considered to have improved turns with medication while red boxes indicate medication non-improved. (**H**) DBS improved turning highlighted with green boxes and non-improved turning highlighted with red boxes. * indicates *p* <0.05, ** indicates *p*<0.01, and *** indicates *p*<0.001.

To explore pallidal-cortical network changes associated with turning, we examined inter-regional coherence differences between turning and straight-line walking. Compared to straight-line walking, GPi-PMC (turning vs Str: 0.26 vs 0.28, *p*=0.0003) and M1-PMC (turning vs Str: 0.33 vs 0.36, *p*=1.51e-10) showed significant decrease in high beta coherence during turning (Figure 2 C, D). There were also significant decreases in alpha (0.26 vs 0.27, *p*=0.0055), low beta (0.28 vs 0.31, *p*=1.64E-7), and low gamma (0.31 vs 0.32, *p*=0.00014) coherence in M1-PMC during turns. Interestingly, GPi-GPe coherence in low beta was increased during turning compared to straight-line walking (0.36 vs 0.39, *p*=0.018).

Overall, these spectral changes highlighted the association of decreased cortical, both M1 and PMC, beta band power during turning. This motivated us to explore if any kinematic variables were correlated with these neurophysiological differences. We found that across all five subjects, pelvic angular acceleration during turning was negatively correlated with M1 low and high beta power (correlation coefficient range: -0.04 to -0.17, p-value range 2.27E-26 to 0.0010, Supplementary Figure 2, Figure 2E). PMC high beta power was also negatively correlated with pelvic angular acceleration (correlation coefficient range: -0.05 to -0.2, p-value range 1.59E-24 to 0.0076, Supplementary Figure 2).

### Turning performance index quantifies interindividual improvement with medication and DBS

To characterize turn quality, we identified 27 distinct measurements associated with turning performance based on literature review (Figure 3A). ^5–11,13,39–41^ Principal component analyses of the 461 turning trials in all subjects across all medication and stimulation states were used to identify the most salient features (Figure 3B). The first ten principal components explained 88.23% of the variance, and the first two explained 41.4%. Analysis of the turning parameters most correlated with the first two principal components revealed that measures of turning speed, both linear and angular speed, as well as stability were most influential (Figure 3C). Stability was assessed by measuring the position of the COM relative to the base of support, with an illustrative example in Figure 3D showing a turn with stable steps throughout alongside a turn featuring some unstable moments. Angular velocity, stability, and linear speed were then combined into a turning performance index (Figure 3E). Our TPI was correlated with clinical rating of turn quality (*r*=0.659, *p*<0.01, Figure 3F).

To evaluate the effects of medication state or DBS on turn quality, we used our validated TPI to quantitively rate all the turns across different medication and stimulation conditions. Subjects 1-4 had significantly improved turning (all *p* <0.001) ON med compared to OFF med with subject 3 showing the smallest change in TPI comparing ON vs OFF med conditions (1.36 vs 1.9) among patients with medication improved turning. Subject 5 showed worse turning ON med compared to OFF med (TPI: 0.72 vs 0.39, *p*=0.016, Figure 3G). Next, we evaluated the effect of DBS on turn quality. Compared to turn trials with DBS OFF, turning performance worsened for subjects 1, 3, and 4 (*p*<0.001) with DBS ON. Turning performance did not significantly differ for subject 2 (TPI: 1.34 vs 1.25, *p*=0.099). Only subject 5 showed a significant improvement in turning with DBS and medication compared to medication alone (TPI: 0.39 vs 1.57, *p*<0.001, Figure 3H).

### Beta band dynamics are associated with turning performance

Given the variable responses to medication and DBS, we hypothesized that turn quality is correlated with shared neurophysiological features. The three patients with the greatest response to medication (subjects 1, 2, 4) were designated as the medication responders while the subject 3 with the least response to medication and subject 5 with worse turning after medication were placed in the medication non-responder group.

In medication responders, turning in the ON state was characterized by broad suppression of beta power in both the pallidum and cortex, accompanied by reduced cortico-pallidal coherence. By contrast, non-responders showed little change across states. In the GPi, low beta (7.79% to 2.64%, *p*=6.71E-13) and high beta power (1.80% to 1.43%, *p*=0.0035; Figure 4A, B) decreased substantially with medication. A similar pattern was seen in the GPe, where low beta power decreased from 4.16% to 2.33% (*p*=5.63E-9) and high beta from 1.98% to 1.35% (p=3.44E-9; Figure 4B) in the ON state. Cortical changes were most prominent in M1, where high beta power decreased in responders (4.99% to 4.02%, *p*=0.0017), and PMC high beta power increased in non-responders (2.83% to 3.28%, *p*=0.048). Connectivity analyses revealed that in the medication responders, GPe–M1 coherence decreased across low beta (0.27 to 0.25, *p*=0.0028), high beta (0.30 to 0.27, *p*=1.4E-7), and low gamma bands (0.28 to 0.26, *p*=2.88E-6) ON med, while GPi–GPe coherence increased ON med (0.32 to 0.36, *p*=0.0082). Individual subject responses to medication are summarized in Supplementary Figure 3.

**Figure 4.**
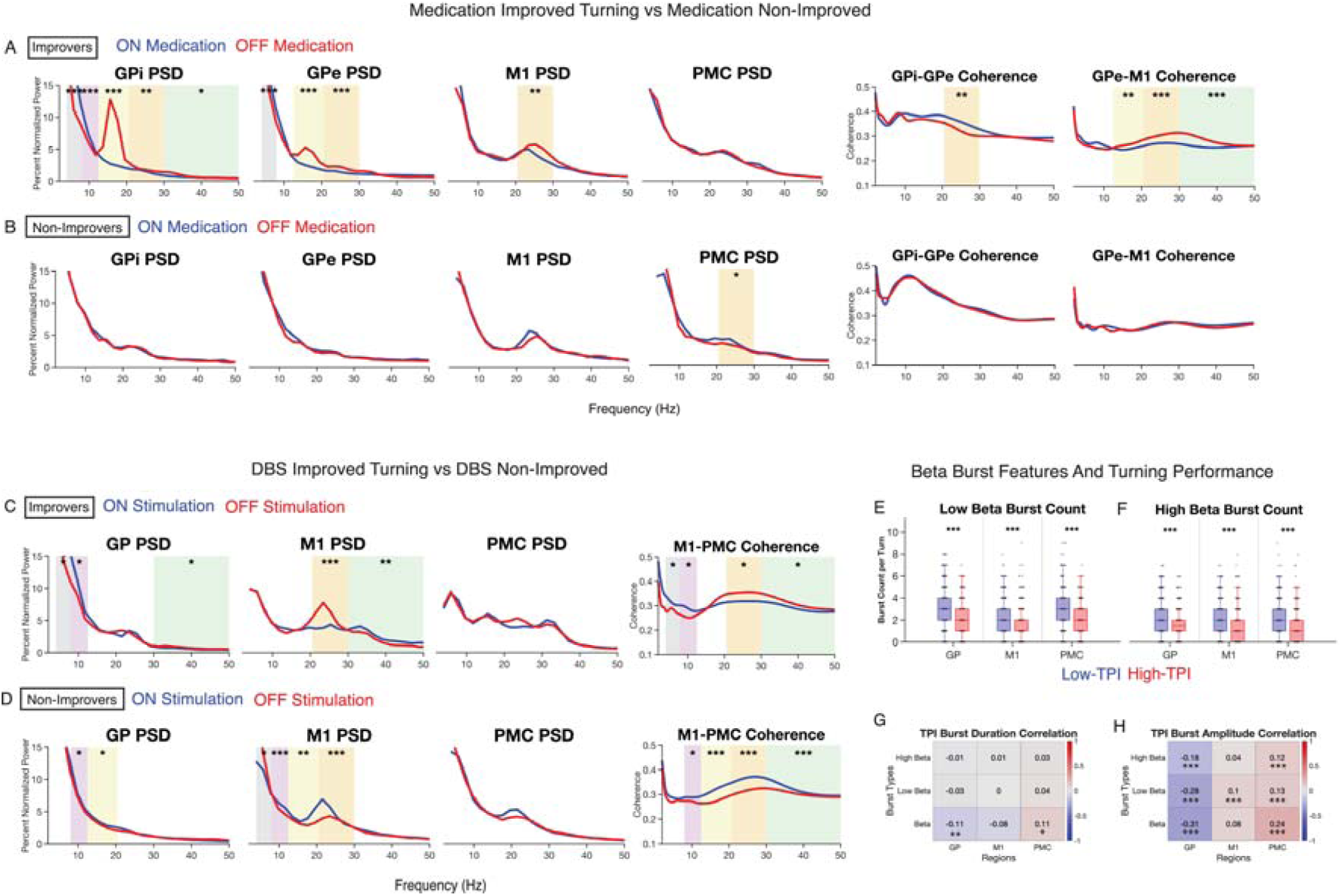
Neural features of medication and stimulation improved turns. (**A**) Normalized power spectral density of turning trials in the OFF medication state and ON medication state. First row represents subjects with medication improved turning and **(B)** second row represents non-improved turning. Globus pallidus internus (GPi), globus pallidus externus (GPe), primary motor cortex (M1), and premotor cortex (PMC) power spectral densities (PSDs) are shown. Inter-regional coherence between medication states in GPi-GPe and M1-PMC are also included. (**C**) Globus pallidus (GP), M1, and PMC PSD curves comparing stimulation ON and medication ON to medication ON alone among subjects with DBS improved turns in the first row and DBS non-improved turns in the **(D)** second row. M1-PMC coherence by stimulation state among these groups. (**E**) Average number of low beta and (**F**) high beta bursts during turns across brain regions in “Low TPI” and “High TPI” trials. (**G**) Beta burst duration and (**H**) amplitude correlated with TPI across all trials. For all panels, * indicates *p* <0.05, ** indicates *p*<0.01, and *** indicates *p*<0.001. Shaded colored regions in spectral power and coherence plots indicate a significant difference in the specified frequency band (theta 4-8 Hz, alpha 8.5-12 Hz, low beta 13-20 Hz, high beta 20.5-30 Hz, and low gamma 30.5-50 Hz).

To examine the neurophysiological changes associated with DBS stimulation, subjects 2 and 5 were classified as responders, as their turning performance improved or did not significantly worsen with DBS, while subjects 1, 3, and 4 were considered non-responders due to worsened turning performance. DBS responders showed cortical beta suppression and reduced cortico-cortical coherence, whereas non-responders frequently exhibited the opposite pattern. In responders, M1 high beta power decreased significantly with stimulation (5.65% to 3.83%, *p*=1.95E-7), accompanied by an increase in low gamma power (1.86% to 2.44%, *p*=0.0062). In contrast, DBS non-responders demonstrated increases in M1 low beta (3.22% to 4.24%, *p*=0.0031) and high beta power (3.61% to 4.50%, *p*=3.91E-5; Figure 4C, D) ON DBS compared to OFF DBS. Pallidal beta power changes were only seen in non-responders, where GP low beta power increased during stimulation (2.88% to 3.27%, *p*=0.045).

Connectivity analyses further highlighted these divergent patterns: in responders, DBS decreased M1–PMC coherence in high beta (0.35 to 0.32, *p*=0.021) and low gamma bands (0.31 to 0.29, *p*=0.021), while non-responders showed marked increases in low beta (0.28 to 0.32, *p*=1.2E-8), high beta (0.32 to 0.37, *p*=1.2E-8), and low gamma coherence (0.30 to 0.32, *p*=0.0042) with stimulation. Individual subject responses to stimulation are summarized in Supplementary Figure 4.

Because our exploration of neural features associated with improved turning underscored the relevance of beta power, we examined beta bursting dynamics, particularly average burst count per turn, burst duration, and burst amplitude across all turns for the low and high beta bands. After dividing all turn trials into “Low TPI” and “High TPI” by the median, we observed significantly more beta bursts per turn across all brain regions in the low TPI group for low beta (GP: 3.07 vs 2.11, *p*=1.56E-12, M1: 2.48 vs 1.73, *p*=1.44E-12, PMC: 2.80 vs 1.83, *p*=3.94E-16, Figure 4E) and high beta (GP: 2.38 vs 1.66, *p*=2.57E-9, M1: 2.22 vs 1.37, *p*=7.50E-15, PMC: 2.29 vs 1.58, *p*=4.26E-10, Figure 4F). Burst duration for the entire beta band was negatively correlated with TPI in the pallidum (*r*= -0.11, *p*=0.0058) but positively correlated in the PMC (*r*=0.11, *p*=0.01; Figure 4G). Burst amplitude negatively correlated with TPI for both low beta (*r*= -0.28, *p*=2.75E-32) and high beta (*r*= -0.18, *p*=1.29E-11; Figure 4H) in the pallidum. The inverse relationship was seen for the cortex. M1 low beta burst amplitude was positively correlated with TPI (*r*=0.1, *p*=0.00085), as was PMC low beta (*r*=0.13, *p*=6.17E-7), and high beta (*r*=0.12, *p*=1.91E-5).

### Machine learning models can reliably identify turns and rate quality from neural spectral features

Finally, we sought to combine our insights from describing the neurophysiologic differences between straight-line walking and turning with our novel grading of turning performance into a machine learning approach to classify and rate turns from neural recordings alone. Because the bidirectional neural interface used in our study supports on-board classification with LDA, we trained LDA models with the top 20 subject specific features for distinguishing between straight-line walking and turning identified through a random forest classifier.

Significantly better than chance AUC model performance was achieved for all five subjects, with AUC ranging from 0.64-0.81, and all subjects showing AUC greater than or equal to 0.7 in at least one hemisphere (Figure 5A). Feature importance was visualized with heatmaps, with an example shown in Figure 5B. Once again, cortical features seemed most salient for distinguishing between straight-line walking and turning.

**Figure 5.**
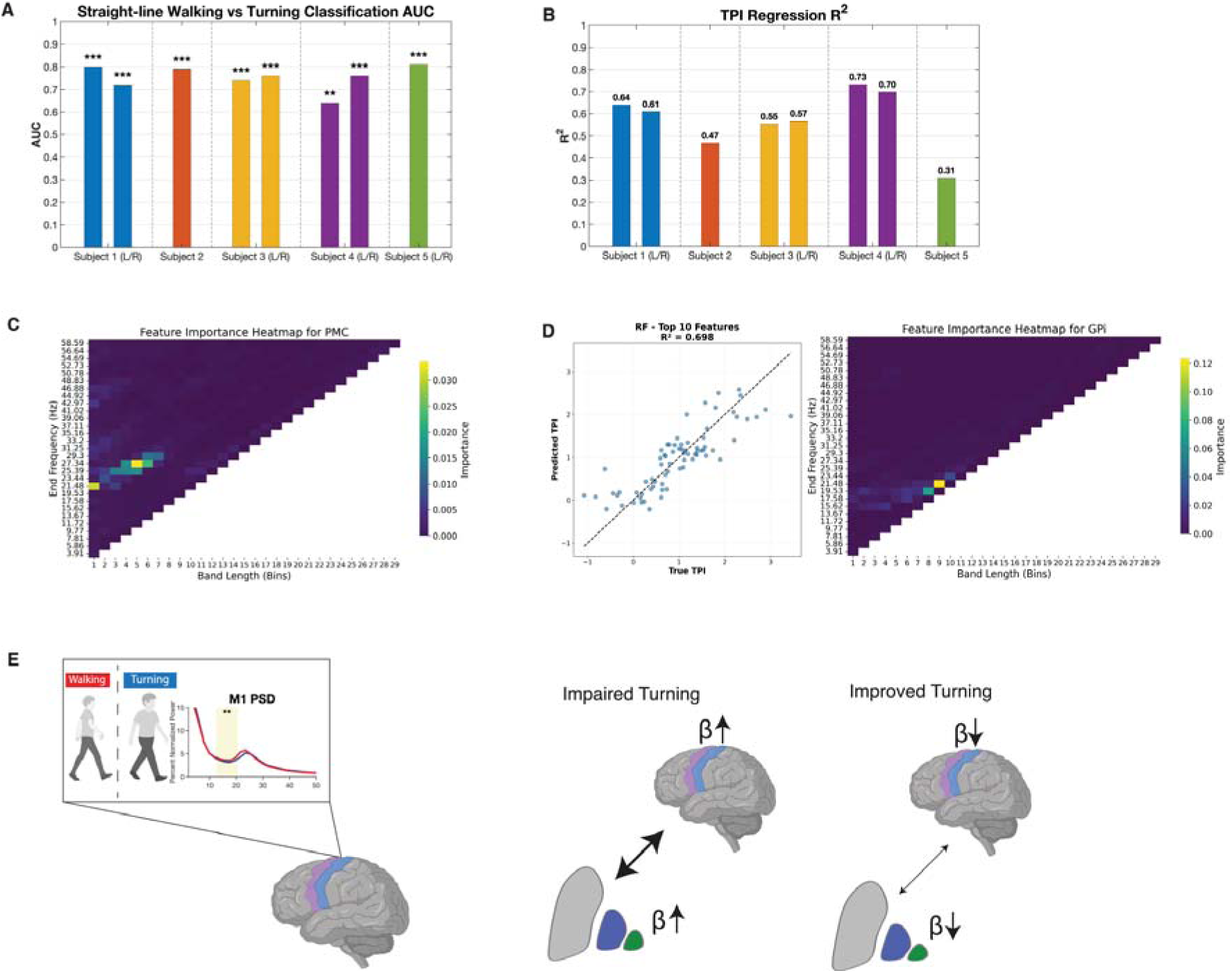
Machine learning models for labeling turns and predicting TPI. (**A**) Cross-validation AUC of linear discriminant analysis (LDA) classifier trained on top 20 subject specific features identified through random forest classifier for distinguishing turns from straight-line walking. (**B**) Example feature importance heatmap for the primary motor cortex (PMC) for a single subject. (**C**) Cross-validation R^2^ values of the top performing turning performance index (TPI) prediction models for each subject. (**D**) Residual plot of predicted versus true TPI along with a heatmap of regression feature importance for one subject. **(E)** Summary schematic showing the use of cortical beta power to separate turning from walking and the cortico-pallidal spectral changes underlying improved and impaired turning.

TPI regression was performed through first ranking all subject specific features by importance, optimizing model hyperparameters among various possible models, and selecting the best performing model. The final cross validation R^2^ values ranged from 0.31 to 0.73 (Figure 5C). Feature importance heatmaps for both classification and regression models, as well as residual plots for the best performing models for each subject are shown in Supplementary Figure 5. The residual plot from subject 4 using the best performing model, a random forest regressor, as well as the feature importance heatmap from the GPi, the brain region with the greatest number of important features among the top 20 features is shown in Figure 5D. Overall, regression analysis identified a mixture of both cortical and pallidal features as playing important roles in predicting TPI (Supplementary Figure 5).

## Discussion

Axial motor symptoms are a uniquely disabling feature of PD due to their ubiquity and poor response to treatment. Turning, in particular, is a routine but highly demanding axial motor behavior, and its impairment is a major cause of falls and morbidity in PD. By combining invasive recordings from pallidal and cortical sites with quantitative kinematic measures, our study reveals circuit signatures that distinguish effective from impaired turning. We show that pathological beta synchronization within cortico-pallidal networks may rigidly lock the motor system into maladaptive states, whereas successful turning requires dynamic desynchronization and flexible network reorganization.

### Axial motor control is enabled by cortical and pallidal beta desynchronization

Turning was consistently associated with suppression of low- and high-beta activity in M1 and PMC, accompanied by reduced cortico-cortical and cortico-pallidal coherence (Figure 5E), differentiating it from straight line walking. In our previous work, we demonstrated through chronic invasive cortical and basal ganglia recordings that straight-line overground walking is associated with low frequency power changes in the subthalamic nucleus and its coherence with M1 time-locked to specific phases of the gait cycle.^28^ These gait-phase associated neural dynamics were also seen in the pallidum, and we can use these biomarkers to distinguish periods of contralateral leg swing during straight-line walking.^50^ In contrast, turning lacked these cyclical oscillations and instead exhibited a global suppression of beta activity across the pallido-cortical network.

Although studies of neurophysiological changes during turns are sparse, we recently reported pallidal and cortical alpha-beta desynchronization during anticipatory postural adjustments in a gait initiation task in PD patients, just prior to the stepping foot leaving the ground.^29^ Additionally, EEG evidence from balance perturbation experiments found that cortical alpha-beta desynchronizations are associated with balance perturbations as well as with voluntary ML and AP sway.^25,51^ Together, these findings suggest that pallidal-cortical beta desynchronization may represent a neurophysiologic correlate of continuous postural transition. In our data, these dynamics correlated with pelvic angular acceleration, suggesting that cortical beta desynchronization may permit the flexibility required to reorient the body axis. By contrast, impaired turns were marked by persistent pallidal and cortical beta bursts, reflecting a temporal rigidity that constrains adaptive control. This supports a mechanistic model in which beta synchronization enforces the motor “status quo,” while its disruption is essential for initiating and executing axial reorientation.^52–58^

### Dopamine and DBS engage parallel pathways for restoring motor flexibility

Our findings also highlight that dopamine and DBS improve turning through distinct yet complementary circuit pathways. In PD, improvements in motor symptoms with levodopa and DBS have been linked to reduced pallidal beta and cortico-pallidal synchronization in the beta band.^59–61^ During simple upper extremity tasks, improved performance on levodopa corresponds to greater cortico-basal ganglia desynchronization at motor onset.^62^ DBS and levodopa can also synergistically improve task performance, as they may preferentially reduce pallidal high and low beta oscillatory power, respectively.^63^

Our findings extend the current understanding of beta dynamics and their relationship with motor performance by showing turning-specific modulation in power and connectivity. Specifically, we show that reductions in low and high beta power both in the GPi and GPe as well as the cortex are associated with improved turning. Medication responders for turning exhibited broad suppression of GPi and GPe beta activity and reduced pallido-cortical coherence, consistent with dopamine restoring basal ganglia throughput. DBS responders for turning, in contrast, showed cortical beta suppression and decreased M1–PMC coherence, underscoring cortical network reorganization as a key therapeutic effect of stimulation. These complementary mechanisms suggest that pallidal and cortical beta represent parallel entry points for restoring motor flexibility during turning.

#### Why DBS may be ineffective for axial symptoms in some patients

Despite the overall efficacy of DBS for appendicular motor symptoms, its impact on axial control is inconsistent.^1,19,21^ In our cohort, some patients demonstrated worsened turning with stimulation, characterized by paradoxical elevations in cortical beta power and M1–PMC coherence compared to ON medication states. Rather than releasing the network from rigid synchronization, stimulation in these individuals may have reinforced maladaptive cortico-cortical coupling, constraining the dynamic flexibility required for turning.

While DBS generally improves motor functions by decreasing basal ganglia and basal ganglia-cortical synchrony, some studies report that DBS can, under certain conditions, increase cortical high beta synchronization.^59,64,65^ In a study comparing resting-state cortical connectivity between DBS on and off states, the authors found that turning DBS on increased high beta and low gamma band synchrony in a cortical circuit spanning the motor, occipitoparietal, middle temporal, and prefrontal cortices.^66^ Additionally, ventral GPi stimulation has been associated with bradykinesia by inhibiting the pedunculopontine nucleus, involved with gait and postural control, or suppressing the motor drive through pallidothalamic fibers.^67–69^ Another possibility is that the elevations in M1 beta power and M1-PMC coherence in the DBS ON state reflect state-specific biomarkers of impaired axial control rather than direct stimulation effects.

Together, these observations underscore that the therapeutic variability of DBS on axial behaviors likely reflects a delicate balance: when stimulation successfully suppresses pathological cortical beta and reduces cortico-cortical coupling, turning improves; when stimulation paradoxically enhances cortical beta synchronization, turning deteriorates. This mechanistic framework suggests that cortical biomarkers may help identify responders versus non-responders and guide individualized programming to avoid maladaptive entrainment of cortical rhythms.

#### Implications for adaptive therapy for axial symptoms

We demonstrate feasibility of using LDA classifiers, the on-board classifier for the neural interface used in our study, to identify turns from straight-line walking using neural signals alone. All subjects achieved an AUC greater than 0.7 in at least one hemisphere, and subject-specific models predicted turning performance with high accuracy. Such approaches could support real-time assessment of turning quality in naturalistic settings and inform adaptive DBS programming tailored to axial performance.

Reliable neural correlates of freezing of gait, impaired turning, and other aspects of gait dysfunction remain an unmet need in PD.^36^ Accurate movement state classification and identification of symptom severity based on biomarkers is a promising strategy for adaptive DBS in PD.^30,36,50,70^ Our integration of pallidal LFP and cortical ECoG recordings is a major strength, enabling both the identification of common network features and the training of individualized models for turn detection and grading. Beyond offline analysis, this framework could dynamically recognize turning as a distinct movement state and transiently adjust stimulation to a gait-optimized setting to improve or stabilize axial function.

#### Limitations

This study has several limitations. Although our analysis represents a novel approach to investigating intracranial neurophysiology during turns, our sample size is limited to five subjects due to the invasive nature of this surgical study with investigational devices. In addition, patient-specific disease factors may limit generalization to broader PD populations or to healthy controls. Although the TPI showed robust agreement with clinician ratings, validation in larger cohorts will be required to confirm external reproducibility. Furthermore, invasive recordings from larger cohorts of patients would enable us to better delineate broad, universal spectral features of impaired turning from patient specific biomarkers.

## Conclusion

Our findings support a mechanistic model in which sub-optimal turning in PD arises from pathological cortico-pallidal beta synchronization that locks the motor system into inflexible states. Effective turning depends on dynamic suppression of beta power and reduction of pathological coherence, enabling flexible reorganization of motor networks. Dopamine and DBS restore turning through distinct circuit pathways, pallidal versus cortical beta suppression, revealing parallel control points for therapeutic modulation. By delineating the oscillatory and connectivity dynamics that distinguish effective from impaired turning, we establish turning as a model behavior for probing axial dysfunction and as a circuit-level biomarker for adaptive stimulation. These insights lay the groundwork for next-generation neuromodulation strategies that dynamically disrupt pathological beta synchronization to restore flexibility in human motor control.

## Data availability

The data, MATLAB and python scripts used in this study can be made available upon reasonable request to the corresponding author. All patient confidentiality and disclosure standards must be adhered to.

## Acknowledgements

We thank the patients for their participation in this study.

---

**Supplementary Figure 1.**
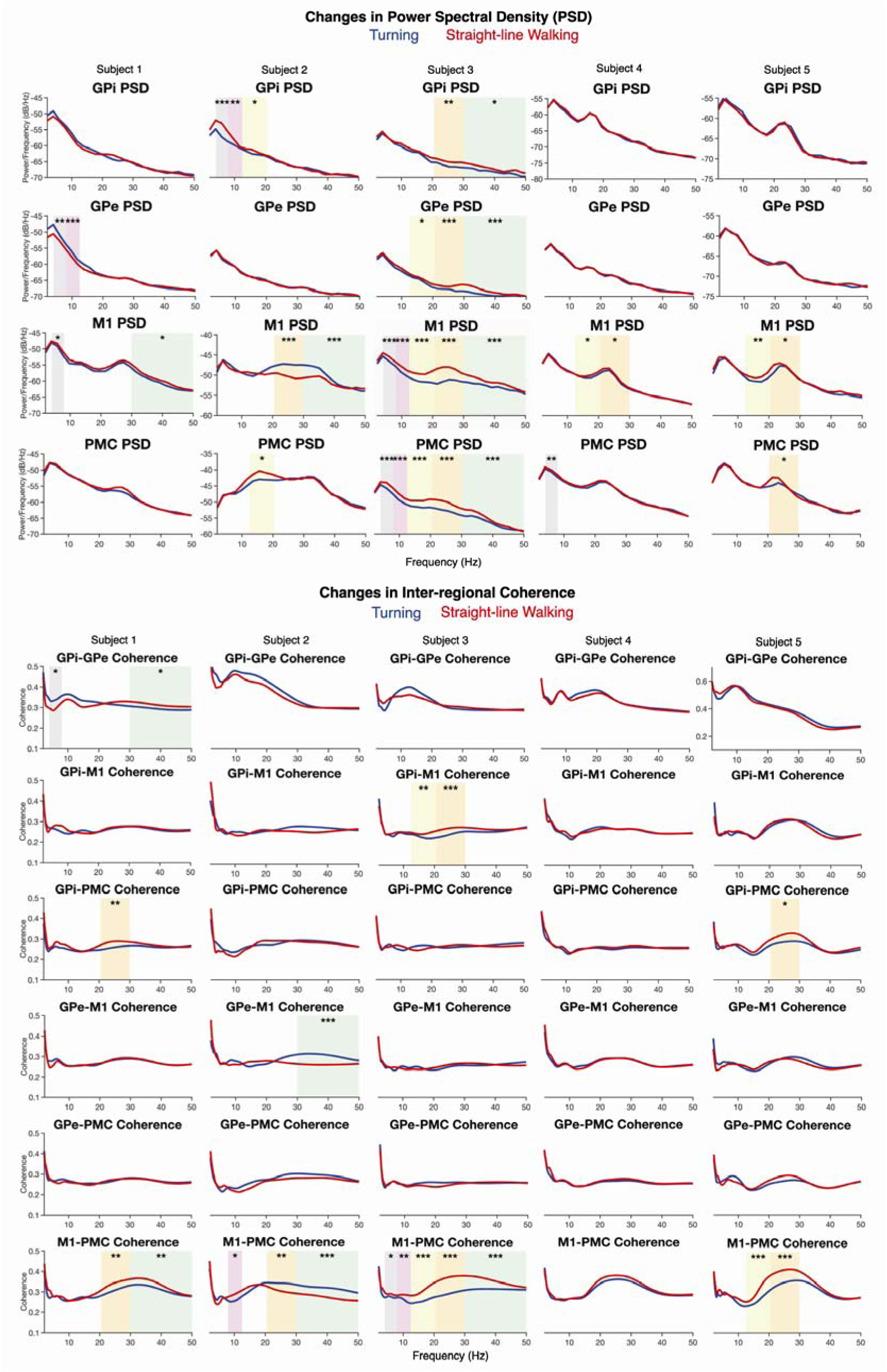
Individual subject neurophysiology of straight-line walking versus turning. Power spectral density (PSD) and inter-regional coherence changes in straight-line walking shown individually for all subjects. For all panels, * indicates *p* <0.05, ** indicates *p*<0.01, and *** indicates *p*<0.001. Shaded colored regions in spectral power and coherence plots indicate a significant difference in the specified frequency band (theta 4-8 Hz, alpha 8.5-12 Hz, low beta 13-20 Hz, high beta 20.5-30 Hz, and low gamma 30.5-50 Hz).

**Supplementary Figure 2.**
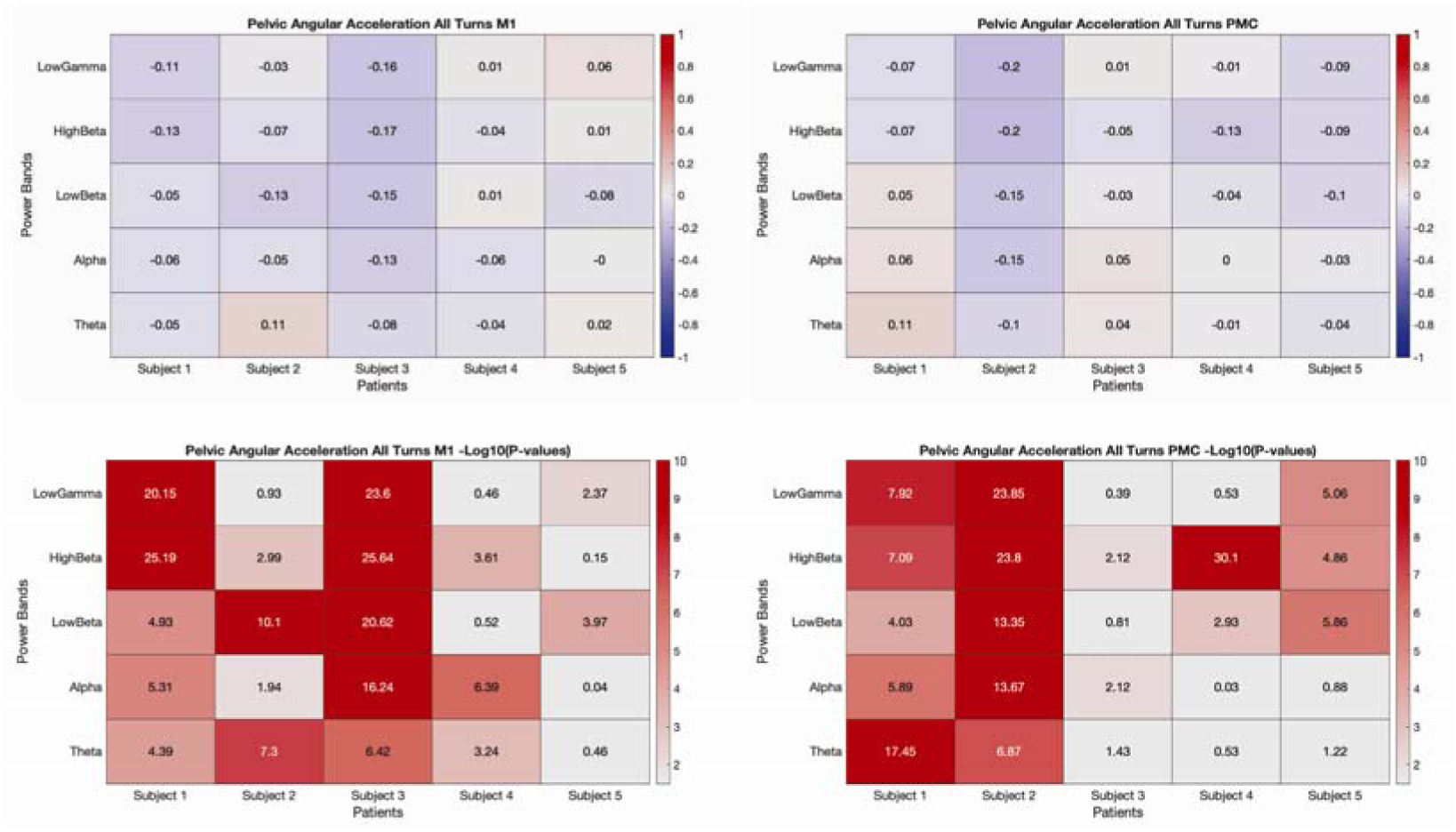
Pelvic angular acceleration correlation with cortical power. Heatmaps showing correlation coefficients (top row) and -log10 p-values (bottom row) for pelvic angular acceleration during all points sampled during turns versus average power in canonical bands for primary motor (M1) and premotor (PMC) cortices. Individual subject, hemisphere, and medication state results are shown in columns of heatmaps.

**Supplementary Figure 3.**
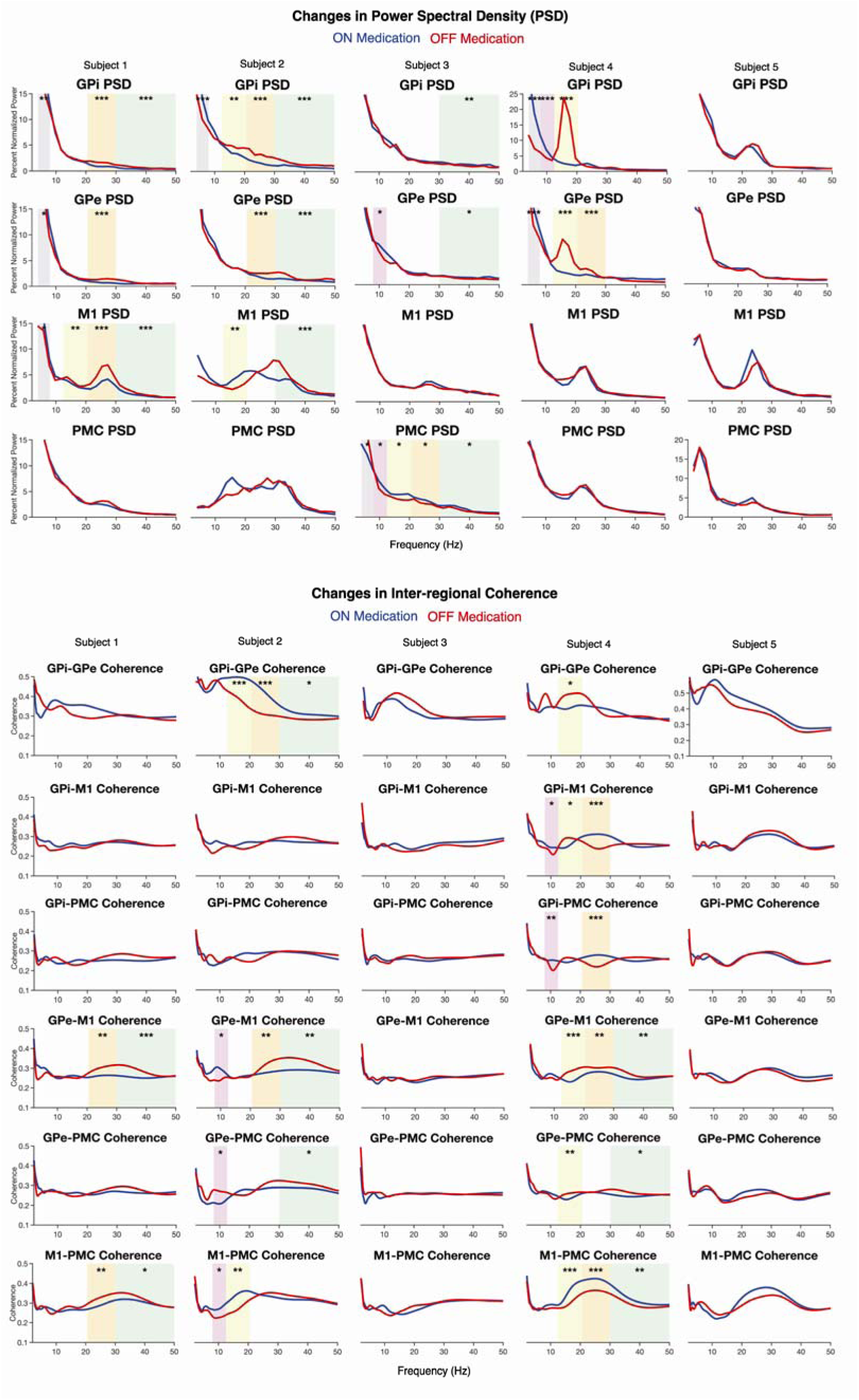
Individual subject neurophysiology of turning response to medication. Power spectral density (PSD) and inter-regional coherence changes during turns in the OFF medication state compared to the ON medication state shown for all subjects. For all panels, * indicates *p* <0.05, ** indicates *p*<0.01, and *** indicates *p*<0.001. Shaded colored regions in spectral power and coherence plots indicate a significant difference in the specified frequency band (theta 4-8 Hz, alpha 8.5-12 Hz, low beta 13-20 Hz, high beta 20.5-30 Hz, and low gamma 30.5-50 Hz).

**Supplementary Figure 4.**
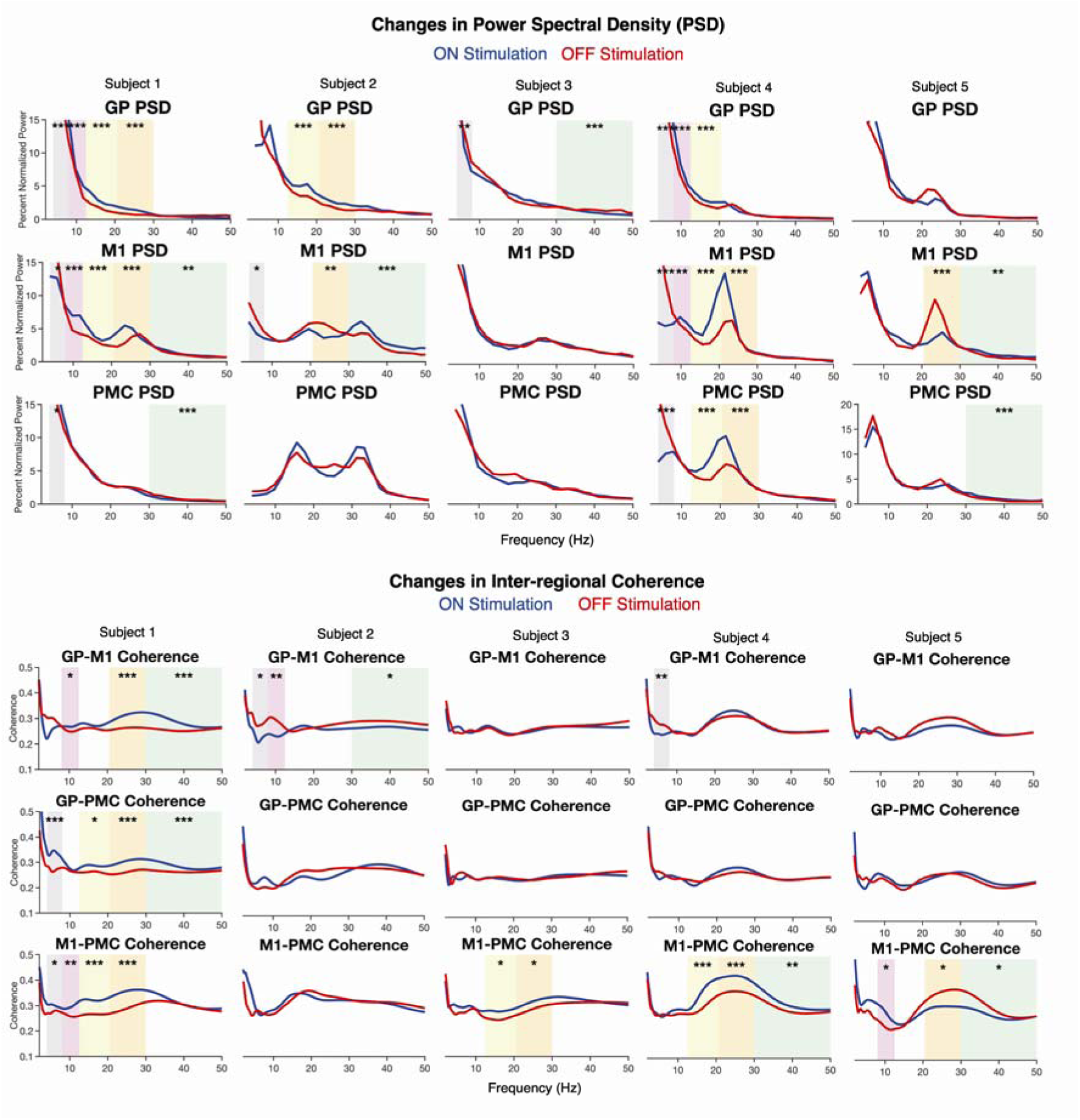
Individual subject neurophysiology of turning response to stimulation. Power spectral density (PSD) and inter-regional coherence changes during turns in the ON stimulation and ON medication state compared to the OFF stimulation and ON medication state shown for all subjects. For all panels, * indicates *p* <0.05, ** indicates *p*<0.01, and *** indicates *p*<0.001. Shaded colored regions in spectral power and coherence plots indicate a significant difference in the specified frequency band (theta 4-8 Hz, alpha 8.5-12 Hz, low beta 13-20 Hz, high beta 20.5-30 Hz, and low gamma 30.5-50 Hz).

**Supplementary Figure 5.**
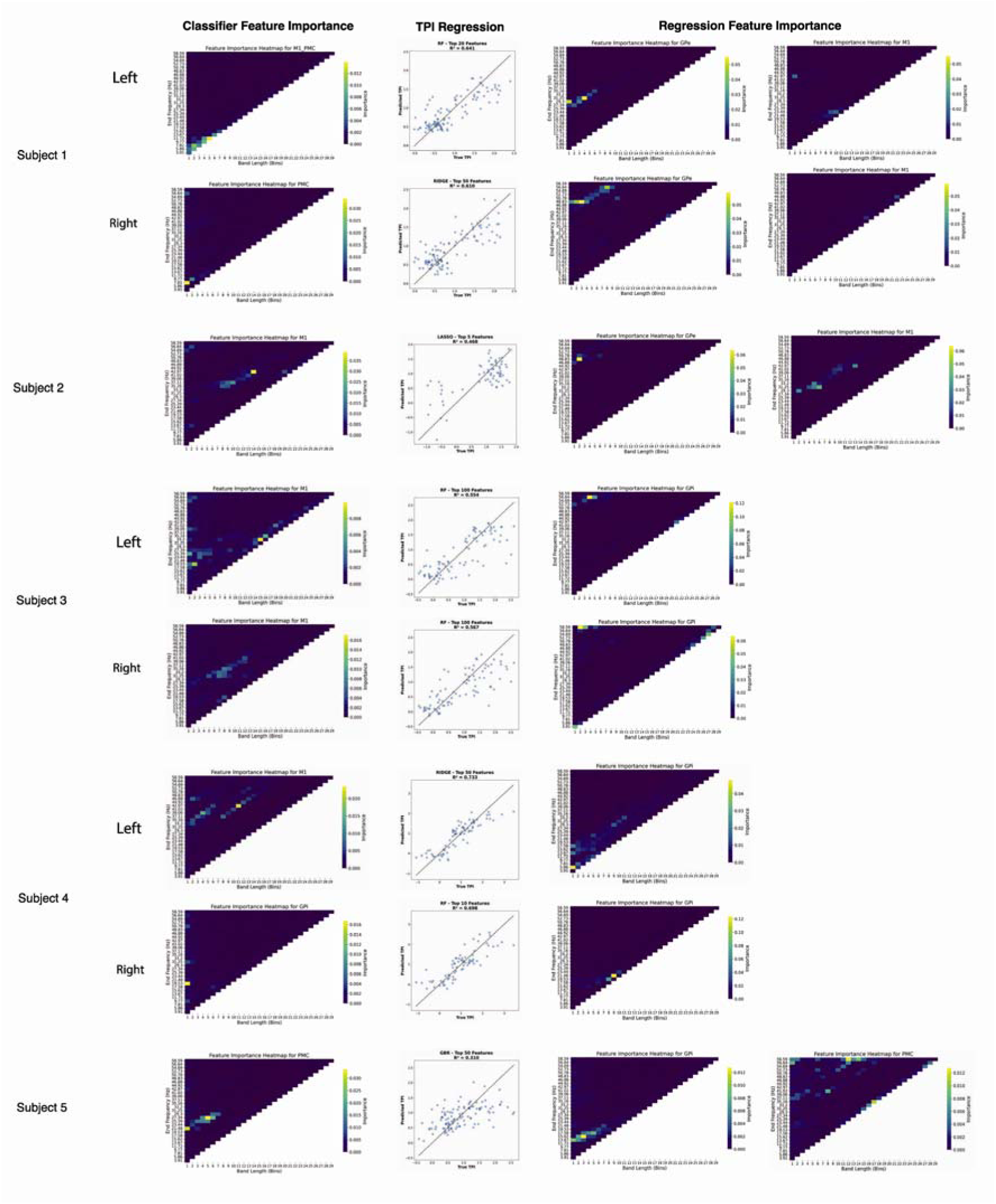
Feature importance and model performance of machine learning models by subject. Feature importance heatmaps from the most important brain regions for the straight-line walking versus turning classification task are shown in the first columns. In the second column, residual plots of the best performing turning performance index (TPI) regression models. Finally, feature importance heatmaps from the most important brain regions for TPI regression.

**Supplementary Table 1.** Subject Demographic and Clinical Characteristics. Demographic and clinical characteristics of included subjects. Age and disease duration are calculated based on date of last trial used for analysis. LEDD (levodopa equivalent daily dose) obtained from latest preoperative data point. MDS-UPRDRS III (Movement Disorders Society-Unified Parkinson’s Disease Rating Scale) and PIGD (postural instability gait disability) as determined by a movement disorders neurologist. *Denotes virtual MDS-UPDRS III, so rigidity and pull-test not evaluated.

| Subject | Age (5-Year Range) | Gender | Disease Duration | LEDD | MDS-UPDRS III OFF MED (PIGD Subscore<sup>1</sup>) | MDS-UPDRS III ON MED (PIGD Subscore<sup>1</sup>) |
| --- | --- | --- | --- | --- | --- | --- |
| 1 | 65-70 | Male | 21 | 2590 | 42 (7) | 25 (5)* |
| 2 | 60-65 | Male | 11 | 1514 | 42 (10)* | 19 (3)* |
| 3 | 57-62 | Female | 8 | 1099 | 57 (3)* | 29 (1)* |
| 4 | 64-69 | Male | 7 | 1025 | 31 (3)* | 17 (2)* |
| 5 | 65-70 | Female | 3 | 618 | 21 (2)* | 9 (2)* |

<sup>1</sup>PIGD Subscore calculated as the sum of MDS-UPDRS III items arising from chair (3.9), gait (3.10), freezing (3.11), postural stability (3.12), posture (3.13)

